# The Impact of Natural Sunlight on Quality of Life and Psychological Well-Being

**DOI:** 10.1101/2025.08.03.25332906

**Authors:** Nino Wessolowski, Michael Rodenkirchen, Svea Mählmann

## Abstract

Sunlight is widely assumed to enhance well-being. However, systematic reviews suggest that the evidence is less extensive and partly inconsistent than commonly assumed.

In this study, *n* = 120 predominantly young adults completed an online survey assessing quality of life (WHOQOL-BREF) and well-being (FAHW). Participants additionally reported how much time they had spent outdoors and at which locations during the preceding 14 days. These self-reports were matched with local and temporal data from the German Weather Service to estimate each participant’s individual sunshine duration.

Correlation analyses revealed significant but small associations between sunshine duration and well-being, as well as between sunshine duration and three of the four quality-of-life domains. The findings support the assumed relationship between sunshine duration, quality of life, and well-being, while also demonstrating a novel approach to determine individual sunshine duration. Future studies should validate this method with larger and more diverse samples. Although increased sun exposure is associated with a higher risk of skin cancer, this aspect was beyond the scope of the present study. The proposed approach may also be applied to studies on artificial light exposure to control for participants’ natural daylight exposure (light history).

## Introduction

A sunny day spent outdoors is commonly perceived as beneficial, as fresh air, brightness, and physical activity are subjectively associated with enhanced well-being. Insufficient exposure to natural daylight has been linked to vitamin D deficiency, impaired bone health, disrupted circadian rhythm and sleep disturbances, decreased well-being [1,2], lower life satisfaction, and reduced quality of life [1]. These associations are becoming increasingly relevant in the context of ongoing digitalization and the growing prevalence of remote work, as people spend less time outdoors. Even before the COVID-19 pandemic, more than 90% of people’s time was already spent indoors [3,4]

Although being indoors does not necessarily preclude exposure to natural daylight, outdoor environments still provide the highest light intensities, despite architectural efforts to integrate glass facades, atria, light wells, or artificial daylight solutions [5]. Increased sun exposure is associated with a higher risk of skin cancer [5]; however, this aspect was beyond the scope of the present study. Previous research on the relationship between sunlight exposure and well-being has predominantly relied on self-reports and questionnaires.

The present study aimed to quantify this relationship more objectively by combining publicly available meteorological data on solar radiation with self-reported time spent outdoors. Specifically, we investigated whether this novel method could demonstrate an association between sunlight exposure and well-being in young, healthy adults in Germany. The findings may inform the development of simple and accessible approaches to assessing individual daylight exposure and its potential effects.

To date, methods for assessing sunlight exposure have ranged from self-report questionnaires, which are economical but prone to recall bias and imprecision [6], to wearable devices such as light-sensing glasses [7], which are limited to small samples due to high costs. More elaborate approaches include analyses of solar radiation intensity using NASA satellite data [8], which remain largely inaccessible to most researchers.

Even when sunlight exposure is not the primary independent variable, its measurement for control purposes is relevant in studies investigating artificial light exposure, as natural daylight constitutes a confounding factor and is often referred to as participants’ *light history* [5,9–12].

### 1.1 Literature Review

The systematic literature search focused on two aspects: (1) studies examining the effects of sunlight on well-being and health and (2) how sunlight exposure was operationalized in these studies. In April 2025, the following databases were searched: APA PsycInfo, APA PsycArticles, Psychology and Behavioral Sciences Collection, CINAHL Complete, Nation Archive (DFG), The New Republic Archive (DFG), Regional Business News, eBook Collection (EBSCOhost), Business Source Premier, MEDLINE Complete, SPORT, SocIndex. The search syntax used was: TI ((sunlight* OR sunshine* OR daylight* OR Sonnenlicht* OR Sonnenschein OR Tageslicht*) AND (“quality of life” OR “well-being” OR Lebensqualität* OR Wohlbefinden*)) This search yielded 6 results, which are discussed in more detail in the literature review section.

### 1.2 Theoretical Background

The effects of natural daylight on mood can primarily be explained by three well-established mechanisms: biological pathways (via vitamin D synthesis and circadian rhythm synchronization) and psychological mechanisms (through conditioning).

#### Vitamin D

In most of the current literature, the effect of sunlight on well-being and quality of life is attributed to biological processes. Exposure to sunlight on the skin leads to the production of vitamin D3 [13]. It is assumed that a change in the vitamin D3 level causes a change in the serotonin level in the brain; for example, a highly significant positive affect and a significant reduction in negative affect are shown after the administration of vitamin D3 [14]. This correlation seems to be confirmed by the observation that exposure to bright light causes a rapid increase in serotonin levels in the brain; however, it is unknown to what extent the retinal uptake of light influences this process [15]. The increased serotonin balance can be explained by increased serotonin synthesis [16]. These assumptions are confirmed by the fact that the serotonin concentration in the neck veins correlates positively with sunlight to which people are exposed [17]. In the central nervous system alone, serotonin is associated with appetite, pain, emotional, social and sexual behaviour, temperature regulation and mood, in addition to the circadian rhythm [18].

#### Circadian rhythm

In addition to the rods and cones of the retina, a third type of receptor has been identified that is particularly sensitive to daylight with a high blue component: the so-called intrinsically photosensitive retinal ganglion cells (ipRGCs) [19]. These receptors cover the retina like a spiderweb and are directly connected to the suprachiasmatic nucleus via the retinohypothalamic tract. This nucleus serves as the primary internal clock for all circadian bodily functions and regulates them through the hormones melatonin and cortisol, as well as the cryptochrome proteins CRY and PER [10].

Melatonin is produced in the pineal gland from the precursor serotonin and is inhibited by light exposure. Cortisol is regulated indirectly via the release of adrenocorticotropic hormone (ACTH), with its rhythm roughly opposing the pattern of melatonin secretion.

Daylight suppresses melatonin levels and promotes its concentrated release (melatonin rebound) in its absence during the evening. The precursor of the sleep hormone melatonin is the “happiness hormone” serotonin, whose concentration increases when melatonin levels are low. Although the so-called melatonin hypothesis in relation to depressive disorders has been refuted, it is still debated to what extent light can improve mood in healthy and depressed individuals—for example, through increased serotonin synthesis or a well-synchronized circadian rhythm and thus a stable internal clock [10].

Moreover, it has been observed that light, in addition to influencing the circadian rhythm, also has direct effects on alertness, cognitive performance and affective responses [5,20–22].

#### Psychological effects

Light may exert psychological effects through mechanisms of classical conditioning [23]. An example of such conditioning in relation to light could be the association of bright sunlight with high activity levels and positive mood, in contrast to inactivity and negative mood during darker daylight conditions. Most people share this learning experience and are accordingly conditioned. In this way, cultural differences in lighting preferences may also be explained [5,12]. However, some individuals may not have had these experiences or may have had different ones, which could account for interindividual differences in responses to light. Knez, [24,25] for example, demonstrated associations between lighting, mood and attention.

### 1.3 Current State of Research

The 6 studies identified through the systematic literature review are summarized in Table 1 and described in the following section.

**Table 1:** Literatur Review.

| Study | Aim | Design | Sample n | Operationalization | Results |
| --- | --- | --- | --- | --- | --- |
| Buscha, F. (2016). Does Daily Sunshine Make You Happy? [26] | Investigation of the effects of various weather parameters, including sunshine, on individual behavior and subjective well-being | n = 10,300 respondents aged 16 years and older | Representative panel study using local British weather data (1991–2008) | Weather data: 226 British weather stations from the British Atmospheric Data Centre; outcomes: well-being and health | No significant or meaningful effects of weather parameters on individual measures of subjective well-being |
| Caldwell, K., Fernandez, R., Traynor, V., & Perrin, C. (2014). Effects of spending time outdoors in daylight on the psychosocial well-being of older people and their family carers: A systematic review. [27] | Review of the effects of outdoor exposure to daylight on the physical health and psychosocial well-being of older adults and informal caregivers | N = 13 studies including adults aged 55 years and older | Summary of experimental and epidemiological study designs published between 1979 and 2013 | Subjective as well as objective outcome variables related to psychosocial well-being, including biomarkers (melatonin, serotonin and vitamin D levels), as well as participants' sleep behavior and physical activity/performance | Narrative presentation of results: Across the four studies examining the effects of daylight on behavioral disorders, no evidence was found for an influence of daylight on physical or verbal aggression. However, some findings suggested that longer time spent outdoors was associated with fewer depressive symptoms. More frequent outdoor exposure over the course of one year was linked to better cognitive maintenance |
| Genuis, S. J. (2006). Keeping your sunny side up. How sunlight affects health and well-being. [28] | Systematic review on the impact of sunlight on human well-being and health | Based on 28 references | Narrative review | Narrative summary of study findings | Sunlight may alleviate depression, pain and vitamin D deficiency. Studies show that sunlight exposure increases serotonin production, thereby enhancing well-being. |
| Korman, M., Tkachev, V., Reis, C., Komada, Y., Kitamura, S., Gubin, D., Kumar, V., & Roenneberg, T. (2022). Outdoor daylight exposure and longer sleep promote wellbeing under COVID-19 mandated restrictions. [29] | Examination of changes in well-being and their association with daylight exposure outdoors, taking into account sleep–wake patterns during the COVID-19 pandemic | N = 7,517 individuals from 40 countries (68.2% female) | Anonymous online survey | 40–45 items assessing daily behavior and lifestyle, including questions on individual sleep behavior and sleep quality and subjective well-being | Social restrictions led to marked changes in daily routines and outdoor behavior (daylight exposure). During the social restrictions, average sleep duration increased by approximately 15 minutes ($p < 0.001$), with a rise of 26 minutes on workdays ($p < 0.001$). Higher daylight exposure, longer sleep duration and reduced social time pressure (e.g., home office, no alarm clock use) were associated with higher levels of subjective well-being, likely due to a smaller discrepancy in social jetlag. |
| Liu, S., Zhang, X., & Zhao, C. (2025). Happiness in the sky: The effect of sunshine exposure on subjective well-being. [30] | Analysis of the influence of sunshine exposure on two dimensions of subjective well-being (“cognitive” and “affective”) | N = 29,582 respondents, M = 47.35 years (SD = 16.24) | Longitudinal study: China Family Panel Studies (CFPS), a nationally representative survey conducted by the Institute of Social Science Survey at Peking University, based on five data waves (2010, 2012, 2014, 2016 and 2018) | Cognitive well-being, self-reported depressive symptoms, sunshine duration | By leveraging intraindividual variation in sunlight exposure over an eight-year period, the study demonstrated a linear association between the duration of sunshine on the interview day and life satisfaction. |
| Swanson, V., Sharpe, T., Porteous, C., Hunter, C., & Shearer, D. (2016). Indoor annual sunlight opportunity in domestic dwellings may predict well-being in urban residents in Scotland. [31] | Study on the impact of annual sunlight exposure in residential indoor spaces on the mental well-being and health of urban dwellers in Scotland | N = 40 residents (aged 18 and older) living in high-rise buildings in Glasgow, Scotland, classified as a socioeconomically disadvantaged area; age range: 41–50 years | Cross-sectional study with behavioral observation | Daily sunshine duration (UK Met Office); additional covariates controlled (e.g., window orientation); outcomes: psychological well-being and physical health | The relationship between annual sunshine exposure and mental well-being was positive and statistically significant. |

Buscha [26] noted that the relationship between weather and well-being had been less extensively investigated than previously assumed. By linking daily weather data with the British Household Panel Survey, the study assessed whether self-reported measures of well-being were associated with temperature, sunshine duration, precipitation and wind speed. The analysis was based on data from 10,300 individuals in the UK. However, only small and statistically non-significant associations were found.

A systematic review by Caldwell et al. [27] examined the effects of outdoor exposure to daylight on the psychosocial well-being of older adults (55 years and older) and their informal caregivers. Thirteen studies were included, some of which reported positive effects on depressive symptoms, cognitive functioning, social participation and quality of life in older adults. For caregivers, however, no conclusive results were found. Due to high methodological heterogeneity, a meta-analysis was not feasible. The authors emphasize the need for further high-quality studies, particularly focusing on psychosocial outcomes in informal caregivers. Genius [28], in his review, highlighted that sunlight is increasingly recognized as a health factor that may help alleviate depression, pain and vitamin D deficiency. Studies have demonstrated that sunlight exposure enhances serotonin production and thus contributes to improved subjective well-being—an effect especially pronounced in cases of seasonal affective disorder. While sunlight had historically been used for therapeutic purposes, it has more recently been avoided due to skin cancer risks. However, newer epidemiological data challenge this view, suggesting that individuals with predominantly indoor lifestyles are particularly at risk. The author advocates for a balanced approach to sunlight exposure—both in medical contexts and in everyday health promotion.

Korman et al. [29] investigated the relationship between subjective well-being, outdoor daylight exposure and sleep-wake behavior during pandemic-related social restrictions as part of the internet-based Global Chrono Corona Survey (n = 7,517; conducted in ten languages). The results clearly indicate that reduced exposure to daylight was associated with significant impairments in various aspects of well-being—particularly sleep quality, quality of life, physical activity and screen time. The authors argue for a causal positive effect of outdoor daylight exposure on subjective well-being.

Liu et al. [30] analyzed data from the China Family Panel Studies (CFPS), a nationally representative longitudinal study conducted across five survey waves (2010–2018; N = 29,582; M = 47.35 years; SD = 16.24). They examined the influence of sunshine duration on two dimensions of subjective well-being (cognitive and affective). Intraindividual fluctuations over the eight-year period revealed a linear relationship between sunshine duration on the day of the interview and reported life satisfaction.

Swanson et al. [31] explored the effect of annual indoor solar radiation on mental well-being and health in urban residents of Scotland (N = 40, aged 41–50), living in high-rise buildings located in a socioeconomically disadvantaged area of Glasgow. This cross-sectional study, which included behavioral observations, incorporated daily sunshine duration data from the UK Met Office, along with additional variables such as window orientation. The relationship between annual solar radiation and psychological well-being was found to be positive and statistically significant.

Overall, the evidence base identified through the systematic literature search regarding the relationship between light and well-being appears less robust in terms of the number of reliable studies than initially expected. This finding aligns with the assessment by Buscha [26], who also pointed to the limited scientific engagement with this topic to date.

Assessment of sunlight exposure: The methods used to assess individual sunlight exposure varied considerably across studies. Self-report questionnaires provide a coarse estimate but are methodologically questionable due to known limitations such as recall bias and low accuracy. Alternatively, some studies—such as those by Buscha and Wessolowski—used weather data, often relying on generalized regional data for all participants, which limits the precision of individual-level analyses.

Technologically based approaches, such as the one used by Guidolin et al. [7], offer a more precise measurement. This study utilized combined light sensors placed at three body locations: near the cornea (mounted on glasses), as a badge worn on the chest and as a wristwatch-like device. This method enabled the identification of country-specific differences in individual light exposure across Germany, Ghana, the Netherlands, Spain and Sweden.

While such technology-based methods provide higher accuracy, they are often feasible only in smaller samples due to cost and logistical constraints. A more practical yet differentiated approach could involve assigning weather data to individual participants, thus enabling larger sample sizes while improving estimates of individual exposure.

### 1.3 Research Question and Aim of the Present Study

Against this background, the research question of the present study is: Can the association between outdoor sunlight exposure, mood and quality of life be detected using the applied method of individual weather data assignment? The aim of this study is to provide further evidence for the hypothesized relationships and to test the presented method for assessing sunlight exposure.

## Materials and Methods

### 2.1 Design

The data were collected as part of an online survey using Unipark software between March 21, 2021 and August 8, 2021. The participation link was distributed across Germany via a snowball sampling approach through the social networks of the participating students.

### 2.2 Variables and operationalization

#### 2.2.1 Independed Variable Sunshine duration

To determine the reported hours of sunshine, respondents were asked where, when and for how long they spent time outdoors. This information was matched with climate data from the Deutscher Wetterdienst (DWD), which provided local sunshine duration with hourly resolution via the Climate Data Center.

The following formula was used to calculate the hours of sunshine for each participant. For each of the 14 study days (i) and for each of the four possible entries per day (j), a time period (k) was defined that represented the duration the participant spent outdoors (a), adjusted for the time interval during which the sun was actually shining in the respective region (b).

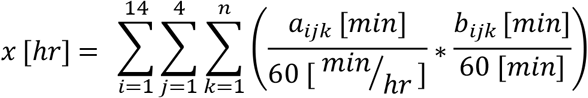

*x:*Sunshine duration of the test subject [in hours]

*i:i* ‘th day of the study

*j:j’*th input field in the question items on behaviour

*k:k’*th hour started of the determined time interval

*n:*Number of full hours started in the analysed time interval

*a:*Length of time spent outdoors in period k [in minutes]

*b:*Sunshine in the subject’s region in time period k [in minutes].

#### 2.2.2 Depended Variable Well-being

The Questionnaire on Habitual Well-Being (FAHW) assesses general well-being by measuring both well-being and dissatisfaction across physical, psychological, and social aspects [32]. The questionnaire consists of twelve items, each answered on a five-point scale, and an overall score can be calculated.

#### 2.2.2 Depended Variable Quality of life

Health-related quality of life comprises physical, mental and social well-being and the ability to function. The “World Health Organization Quality of Life” working group developed the WHOQOL-100 instrument for recording subjective quality of life, from which the WHOQOL-BREF [33] short form was ultimately derived.

The WHOQOL-BREF inventory comprises 26 items, which use a five-point Likert scale to assess the dimensions of physical well-being, psychological well-being, social relationships and environment. Two questions are used for an overall assessment of quality of life.

### 2.3 Sampling approach

Because no suitable empirical reference values were available from previous studies, a medium effect size was assumed for the relationship between sunshine, quality of life and well-being. Statistical power was set to 90 % with a one-tailed α of ≤ 0.05. Based on these parameters, G*Power [34] indicated that a sample size of n = 72 would be required to attain statistical significance.

### 2.4 Data collection

The survey began with the research intention and a few words of welcome from the research team. After informing the participants about the voluntary nature of the anonymous survey and providing legal information, demographic data were collected. This was followed by the FAHW and WHOQOL-BREF questionnaire and the recording of the length of stay experienced outside the home.

### 2.5 Evaluation procedure

The raw data was examined in detail and test subjects who stopped answering prematurely, answered unrealistically quickly or gave implausible answers were excluded on a case-by-case basis, as were test subjects whose locations were not available in the DWD data set.

The sample was described using descriptive statistics, including sample size, range, mean and the corresponding standard error. Further analysis of sunshine duration was then conducted. First, the region entered in the initial input field was verified and the corresponding data set from the German Weather Service (DWD) was selected. Next, the date and time period specified by the respondent for that region were identified. Based on this information, the relevant time window was located within the weather data, allowing the extraction of hourly sunshine values.

To refine the time specification, a mathematical function was applied to determine the exact duration of stay in minutes. As outlined in the function, the sum of all individual hourly values was calculated following the data extraction.

### 2.5 Ethics

The experiment was conducted in accordance with the Declaration of Helsinki and was approved by the Ethics Committee of Medical School Hamburg (MSH-2022/151).

### 2.5 Statistics

To examine the relationship between the predominantly ordinally scaled variables, a Spearman rank correlation analysis was conducted. The analysis was performed using JASP. The classification of effect sizes was based on Cohen [35].

## Results

### 3.1 Sample

A total of *n* = 120 participants were included in the sample. Most of them were young (M = 25.6 years, SD = 9.19) and female students (see Table 2). The optimal sample size was achieved (see Section 2.3, Sampling Approach).

**Table 2:** Basic characteristics.

|  | *n* | % |
| --- | --- | --- |
| Gender |  |  |
| female | 101 | 84.2 |
| male | 17 | 14.2 |
| diverse | 2 | 1.7 |
| Residence <sup>a</sup> |  |  |
| northern germany | 92 | 76.7 |
| central germany | 25 | 20.8 |
| southern germany | 3 | 2.5 |

*Note. N = 120 participants had a mean age of 25.6 years (SD = 9.188). a For the regional classification, the federal states were grouped into the categories North (Schleswig-Holstein, Hamburg, Lower Saxony, Bremen), Central (North Rhine-Westphalia, Hesse, Rhineland-Palatinate, Saarland, Berlin), and South (Bavaria, Baden-Württemberg).*

### 3.2 Results

A descriptive overview of the variables, including mean, standard deviation, minimum, and maximum values, is provided in Table 3. According to Cohen’s [35] interpretation of Spearman’s r, a significant small correlation was found between sunshine duration and well-being. Three additional significant small correlations were observed between sunshine duration and three of the four quality-of-life domains: physical health, mental health, and environment. No significant correlation was found between sunshine duration and social relationships. In summary, four of the five constructs examined showed significant small correlations with sunshine duration (see Table 4).

**Table 3:**
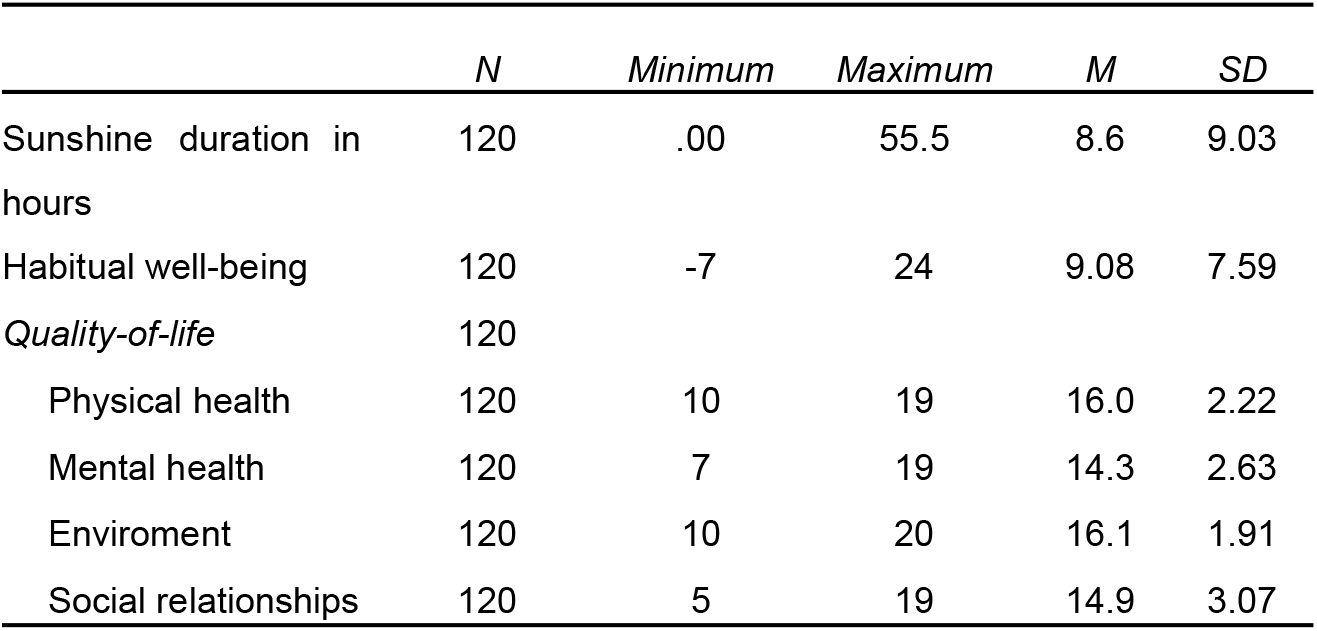
Descriptives.

**Table 4:** Correlation with the variable sunshine duration.

|  | 1 | 2 | 3 | 4 | 5 | 6 |
| --- | --- | --- | --- | --- | --- | --- |
| 1. Sunshine duration in hours |  |  |  |  |  |  |
| 2. Habitual well-being | 0.27** |  |  |  |  |  |
| Quality-of-life |  |  |  |  |  |  |
| 3. Physical health | 0.23* | 0.72*** |  |  |  |  |
| 4. Mental health | 0.23** | 0.73*** | 0.65*** |  |  |  |
| 5. Enviroment | 0.20* | 0.55*** | 0.54*** | 0.54*** |  |  |
| 6.Social relation-ships | -0.02 | 0.39*** | 0.31*** | 0.41*** | 0.10 |  |

Note. *r* = Spearman's rho correlation.

## Discussion

### 4.1 Main Results

The hypothesized associations between sunshine, quality of life and well-being were confirmed. Notably, this study was very easy to conduct as an online survey. No technical effort or intrusion into participants’ privacy was required, such as permanently wearing a light sensor or tracking their exact location.

The results indicate a significant small correlation between well-being and sunshine duration. The study further demonstrates that sunshine duration is significantly and positively associated, with a small effect size, with the domains of physical health, mental health and environmental perception of quality of life. These findings support the use of individually determined sunshine duration data provided by weather services for investigating similar hypotheses.

### 4.2 Limitations

One limitation of the results is that only the exposure to sunshine outdoors was considered and natural light in buildings and artificial light were not taken into account. The values were also approximated using mean values from weather stations and not recorded individually. Thus, the data results from a consideration of the locations of the test persons and the respective measurement data of the DWD. Data is only available for large cities and airports, so the test subjects were assigned to the nearest possible measuring point. In addition, the data used only has an hourly resolution of sunshine duration. However, if intra-individual measurement is ruled out, the use of DWD data appears to be the most economical alternative.

A further limitation is the subjects’ ability to remember the two-week period in question. The recency effect suggests that the data becomes increasingly blurred as the time lag increases. Cognitive distortions can be minimised in future by encouraging the test subjects to create daily documentation for the study period. In the current study, this option, as well as a longitudinal design, was ruled out for economic reasons.

Despite a detailed description and explanation of the task and an example, not all test subjects were able to correctly interpret the question about their whereabouts. In order to avoid having to exclude test subjects in future, the postcode can be collected in addition to the location. This offers the advantage of more accurate measured values if several sunshine sensors are available in a region.

When interpreting the results, particular attention must be paid to the fact that the subjects were not asked why they were out of the house and whether and to what extent they were on the move. This seems relevant, as the effect of physical activity on well-being and quality of life is well known [36,37]. It would therefore be worthwhile investigating the contribution of exercise in future projects. The exercise parameter was not included in this study, as the time required to complete the survey was already considered critical.

### 4.2 Interpretation of the findings

The results show small but significant positive correlations between sunshine duration and habitual well-being as well as three quality-of-life domains: physical health, mental health and environment. In contrast, no significant correlation was observed for social relationships.

These findings are largely consistent with theoretical expectations. Sunshine exposure is assumed to influence well-being and quality of life through multiple mechanisms: (a) biological effects, such as increased vitamin D synthesis and regulation of the circadian rhythm, which may improve mood, sleep, physical functioning and (b) psychological effects, including positive affective responses to daylight and outdoor environments.

The absence of an association with social relationships is not surprising. Theories explicitly linking sunshine exposure to social well-being are scarce, apart from the assumption that people may spend more time outdoors and engage in social interactions during sunny weather. However, this effect is likely minimal during spring and summer, the periods in which the present study was conducted.

Overall, the observed correlations are in line with much of the existing research, even though meta-analyses and systematic reviews have reported inconsistent findings and a weaker evidence base than previously assumed [26,28]. Nevertheless, the present study contributes additional evidence by demonstrating these associations using a technically simple and standardisable procedure for estimating individual sunshine duration in Germany.

### 4.4 Summary

The study presented here shows that the relationship between sunshine duration, well-being and quality of life could be demonstrated for the first time using a simple and minimally invasive technique. In principle, it is possible to dispense with more complex and invasive techniques such as light meters, as long as the inaccuracies and weaknesses of the method can be tolerated, as described in the methodological critique.

The invasiveness of more precise measurements with light sensors on glasses becomes clear if you imagine, as a participant in a study, wearing glasses with a light measurement device all day without interruption, even in areas such as the bedroom or bathroom. Alternatively, the evaluation of movement data from the mobile phone could be considered, whereby the researchers would prefer the daily and voluntary request of movement data in order to counteract the criticised memory bias of the method presented here on the one hand, but on the other hand to ensure maximum data protection and preservation of the privacy of the participants.

Despite the limitations mentioned, this pilot study with the simplified measurement method shows small but significant correlations. Evidence for a long-suspected and theoretically well-founded correlation can thus be found and presented.

This once again emphasises the relevance of being able to spend time outdoors and should therefore be taken into account, for example, in urban development that aims to design living spaces with positive attributes where residents enjoy spending time. However, it is important to also consider the increased risk of skin cancer associated with sunlight exposure, meaning that spending more time outdoors cannot be unconditionally recommended. The importance of daylight planning—in the design of buildings, whether in private environments, manufacturing facilities, office workplaces, clinics, schools, or public buildings—is likewise acknowledged. This is best ensured by natural daylight, as in this study, or alternatively by artificial daylight when natural light is insufficient.

## Data Availability

The data have been submitted as supplementary material.

## References

1. de Kort Y. Light and Quality of Life. Encyclopedia of Quality of Life and Well-Being Research. Springer, Dordrecht; 2014. pp. 3615–3620. doi:10.1007/978-94-007-0753-5_3961

2. Holick MF. Chapter 2 - A perspective on the beneficial effects of moderate exposure to sunlight: bone health, cancer prevention, mental health and well being. In: Giacomoni PU, editor. Comprehensive Series in Photosciences. Elsevier; 2001. pp. 11–37. doi:10.1016/S1568-461X(01)80037-5

3. Pitarma R, Marques G, Ferreira BR. Monitoring Indoor Air Quality for Enhanced Occupational Health. J Med Syst. 2016;41: 23. doi:10.1007/s10916-016-0667-2

4. Jiang Y, Li K, Tian L, Piedrahita R, Yun X, Mansata O, et al. MAQS: a mobile sensing system for indoor air quality. Proceedings of the 13th international conference on Ubiquitous computing. New York, NY, USA: Association for Computing Machinery; 2011. pp. 493–494. doi:10.1145/2030112.2030187

5. Wessolowski N. Licht, Farbe und Psyche: Theorie, Forschung und praktische Empfehlungen. 2nd ed. Hamburg: epubli; 2024.

6. Wessolowski N. Einführung in die Forschungsmethodik: Wissenschaftliches Arbeiten und Statistik: mit praktischer Durchführung, Prüfungsfragen, Umgang mit Künstlicher Intelligenz & vielem mehr. 1st ed. Berlin: epubli; 2023.

7. Guidolin C, Aerts S, Agbeshie GK, Akuffo KO, Aydin SN, Baeza-Moyano D, et al. Protocol for a prospective, multicentre, cross-sectional cohort study to assess personal light exposure. BMC Public Health. 2024;24: 3285. doi:10.1186/s12889-024-20206-4

8. Kent ST, McClure LA, Crosson WL, Arnett DK, Wadley VG, Sathiakumar N. Effect of sunlight exposure on cognitive function among depressed and non-depressed participants: a REGARDS cross-sectional study. Environ Health. 2009;8: 34. doi:10.1186/1476-069X-8-34

9. Barkmann C, Wessolowski N, Schulte-Markwort M. Applicability and efficacy of variable light in schools. Physiol Behav. 2012;105: 621–627. doi:10.1016/j.physbeh.2011.09.020

10. Wessolowski N. Wirksamkeit von Dynamischem Licht im Schulunterricht. doctoralThesis, Staats-und Universitätsbibliothek Hamburg Carl von Ossietzky. 2014. Available: https://ediss.sub.uni-hamburg.de/handle/ediss/5418

11. Wessolowski N, Koenig H, Schulte-Markwort M, Barkmann C. The effect of variable light on the fidgetiness and social behavior of pupils in school. J Environ Psychol. 2014;39: 101–108. doi:10.1016/j.jenvp.2014.05.001

12. Wessolowski N, Rahim RJ. Is executive attention affected by environmental lighting conditions? Steinborn MB, editor. PLOS ONE. 2025;20: e0305998. doi:10.1371/journal.pone.0305998

13. Webb AR, Holick MF. The Role of Sunlight in the Cutaneous Production of Vitamin D3. Annu Rev Nutr. 1988;8: 375–399. doi:10.1146/annurev.nu.08.070188.002111

14. Lansdowne ATG, Provost SC. Vitamin D3 enhances mood in healthy subjects during winter. Psychopharmacology (Berl). 1998;135: 319–323. doi:10.1007/s002130050517

15. aan het Rot M, Benkelfat C, Boivin DB, Young SN. Bright light exposure during acute tryptophan depletion prevents a lowering of mood in mildly seasonal women. Eur Neuropsychopharmacol. 2008;18: 14–23. doi:10.1016/j.euroneuro.2007.05.003

16. aan het Rot M, Moskowitz DS, Young SN. Exposure to bright light is associated with positive social interaction and good mood over short time periods: A naturalistic study in mildly seasonal people. J Psychiatr Res. 2008;42: 311–319. doi:10.1016/j.jpsychires.2006.11.010

17. Lambert G, Reid C, Kaye D, Jennings G, Esler M. Effect of sunlight and season on serotonin turnover in the brain. The Lancet. 2002;360: 1840–1842. doi:10.1016/S0140-6736(02)11737-5

18. Kriegebaum C, Gutknecht L, Schmitt A, Lesch K-P, Reif A. Serotonin Kompakt – Teil 1. Fortschritte Neurol · Psychiatr. 2010;78: 319–331. doi:10.1055/s-0029-1245240

19. Berson DM, Dunn FA, Takao M. Phototransduction by Retinal Ganglion Cells That Set the Circadian Clock. Science. 2002;295: 1070–1073. doi:10.1126/science.1067262

20. Golmohammadi R, Yousefi H, Safarpour Khotbesara N, Nasrolahi A, Kurd N. Effects of Light on Attention and Reaction Time: A Systematic Review. J Res Health Sci. 2021;21: e00529. doi:10.34172/jrhs.2021.66

21. Mu Y-M, Huang X-D, Zhu S, Hu Z-F, So K-F, Ren C-R, et al. Alerting effects of light in healthy individuals: a systematic review and meta-analysis. Neural Regen Res. 2022;17: 1929. doi:10.4103/1673-5374.335141

22. Lasauskaite R, Wüst LN, Schöllhorn I, Richter M, Cajochen C. Non-Image-Forming Effects of Daytime Electric Light Exposure in Humans: A Systematic Review and Meta-Analyses of Physiological, Cognitive, and Subjective Outcomes. LEUKOS. 0: 1–42. doi:10.1080/15502724.2025.2493669

23. Rea M. Light-Much More Than Vision. 2003. Available: https://www.semanticscholar.org/paper/Light-Much-More-Than-Vision-Rea/db8f5ba0be450279bce83054781b513a70fc35ff?p2df

24. Knez I. Effects of indoor lighting on mood and cognition. J Environ Psychol. 1995;15: 39–51. doi:10.1016/0272-4944(95)90013-6

25. Knez I, Enmarker I. Effects of Office Lighting on Mood and Cognitive Performance And A Gender Effect in Work-xRelated Judgment. Environ Behav. 1998;30: 553–567. doi:10.1177/001391659803000408

26. Buscha F. Does Daily Sunshine Make You Happy? Subjective Measures of Well-Being and the Weather. Manch Sch 1463-6786. 2016;84: 642–663. doi:10.1111/manc.12126

27. Caldwell K, Fernandez R, Traynor V, Perrin C. Effects of spending time outdoors in daylight on the psychosocial well-being of older people and their family carers: a systematic review. JBI Database Syst Rev Implement Rep. 2014;12: 277–320. doi:10.11124/jbisrir-2014-1604

28. Genuis SJ. Keeping your sunny side up. How sunlight affects health and well-being. Can Fam Physician Med Fam Can. 2006;52: 422.

29. Korman M, Tkachev V, Reis C, Komada Y, Kitamura S, Gubin D, et al. Outdoor daylight exposure and longer sleep promote wellbeing under COVID-19 mandated restrictions. J Sleep Res. 2022;31.

30. Liu S, Zhang X, Zhao C. Happiness in the sky: The effect of sunshine exposure on subjective well-being. Biodemography Soc Biol. 2025;70: 67–81. doi:10.1080/19485565.2025.2487977

31. Swanson V, Sharpe T, Porteous C, Hunter C, Shearer D. Indoor annual sunlight opportunity in domestic dwellings may predict well-being in urban residents in Scotland. Ecopsychology. 2016;8: 121–130. doi:10.1089/eco.2015.0059

32. Wydra G. FAHW. Fragebogen zum allgemeinen Wohlbefinden. 2024 [cited 31 Jul 2025]. Available: https://psycharchives.org/en/item/56b52443-5fb4-411e-a016-bb923a4ccab9

33. WHO. German_WHOQOL-BREF. 2025 [cited 31 Jul 2025]. Available: https://www.who.int/nepal/activities/supporting-elimination-of-kala-azar-as-a-public-health-problem/docs/default-source/publishing-policies/whoqol-bref/german-whoqol-bref

34. Faul F, Erdfelder E, Buchner A, Lang A-G. Statistical power analyses using G*Power 3.1: Tests for correlation and regression analyses. Behav Res Methods. 2009;41: 1149–1160. doi:10.3758/brm.41.4.1149

35. Cohen J. Statistical Power Analysis for the Behavioral Sciences. New York, NY: Taylor & Francis; 1988.

36. Anokye NK, Trueman P, Green C, Pavey TG, Taylor RS. Physical activity and health related quality of life. BMC Public Health. 2012;12: 624. doi:10.1186/1471-2458-12-624

37. Fox KR. The influence of physical activity on mental well-being. Public Health Nutr. 1999;2: 411–418. doi:10.1017/S1368980099000567

